# Food Insecurity Among Older Adults with a History of Incarceration

**DOI:** 10.1101/2022.02.08.22270608

**Authors:** Tamara Jordan, Rodlescia Sneed

**Author notes:** **Corresponding Author:** Dr. Tamara Jordan, DO, MPH, MLS; Michigan State University – Division of Public Health; 200 East 1^st^ St., Flint, MI 48502, USA.

## Abstract

**Objectives:** To examine the association between history of incarceration (HOI) and food insecurity (FI) among older adults.

**Methods:** This is a secondary analysis utilizing data from 12,702 respondents aged 51+ who participated in the 2012 and 2014 waves of The Health and Retirement Study. Multiple logistic regression was used to estimate the association between HOI and FI, adjusting for demographic variables using odds ratios (OR) and 95% confidence intervals (CI).

**Results:** In our sample, 12.8% of participants reported FI. Having a HOI increased odds of FI (OR 1.83; 95% CI 1.52-2.21), especially among Blacks (OR 1.78; 95% CI 1.29-2.46) and Whites (OR 2.27; 95% CI 1.74-2.97), but not Hispanics (OR 1.11; 95% CI 0.69-1.77) or other racial/ethnic groups (OR 1.79; 95% CI 0.71-4.52).

**Discussion:** FI is an important issue among older adults with a HOI. Stronger linkages between formerly incarcerated older adults and existing food assistance programs are needed.

## Introduction

Food insecurity (i.e., the inability to always access an appropriate quantity and quality of food to maintain an active and healthy lifestyle) is a growing problem among older adults. In the United States, about one in twelve adults aged 50 and older report being food insecure, which translates to over 9 million older adults (Gunderson & Ziliak, 2021; Ziliak & Gunderson, 2021). Numerous studies suggest that food insecurity among older adults can be attributed to chronic disease burden (Jih et al., 2018), functional limitations (Chang & Hickman, 2018), and residing in multigenerational households (Ziliak & Gunderson, 2016). Food insecurity is an important public health problem, as it is associated with greater risk of malnutrition and numerous chronic health conditions, including diabetes, high blood pressure, and depression (Gunderson & Ziliak, 2015).

Older adults who have been previously incarcerated may be particularly at risk of food insecurity. There is a growing body of evidence suggesting that justice system involvement among older adults is rapidly increasing. From 1993 to 2013, the number of adults aged 55+ incarcerated in state prisons increased by 400%, likely due to an increase in conviction rates among older adults and the use of longer prison sentences as a strategy to promote public safety (Carson & Sabol, 2016). Additionally, there has been an increase in the number of older adults returning to community settings after incarceration, as states attempt to decarcerate older prisoners in an effort to reduce healthcare spending associated with caring for an older population (Williams & Abraldes, 2007). Once released from prison, these older adults often face numerous challenges that increase their risk of food insecurity, including homelessness (Couloute, 2018), as well as unemployment and poverty (Western & Sirois, 2019). Several studies conducted in younger samples suggest that incarceration is associated with increased risk of food insecurity (Testa & Fahmy, 2021; Testa & Jackson, 2019; Testa & Jackson, 2020). These risk factors are likely magnified among older adults with a history of incarceration, as they likely also face age-related barriers to employment (e.g., age discrimination; Harootyan, 2021) and mobility issues (Bjelland et al., 2010), that further increase the risk of food insecurity.

Given that ninety-five percent of incarcerated individuals are eventually released (Hughes & Wilson, 2003), and the rapid increase in justice system involvement among older adults, it is crucial to understand the health-related experiences of formerly incarcerated older adults after they reenter communities. While the association between previous incarceration and food insecurity has been explored in younger adults (Testa & Fahmy, 2021; Testa & Jackson, 2019; Testa & Jackson, 2020), there has been little attention to food insecurity in older samples. Further, few studies have utilized population-based studies to examine this association. The purpose of this current study is to use a population-based sample of adults to examine the association between history of incarceration and food insecurity among community-dwelling older adults.

We were also interested in whether race/ethnicity moderated the association between history of incarceration and food insecurity. There are large racial/ethnic minority disparities in food insecurity. Nineteen percent of non-Hispanic Black households and 15.6% of Hispanic households report food insecurity, compared to only 7.9% of non-Hispanic White households (Coleman-Jensen et al., 2020). Additionally, racial/ethnic minorities are more likely to be incarcerated than non-Hispanic White Americans. While non-Hispanic Blacks make up only 13.4% of the U.S. population (U.S. Census Bureau, 2019), they account for 38.1% of those currently incarcerated (Federal Bureau of Prisons, 2022). Likewise, Hispanics make up 18.5% of the total U.S. population (U.S. Census Bureau, 2019), but account for 30.4% of those currently incarcerated (Federal Bureau of Prisons, 2022). Given these data, we would expect that non-Hispanic Blacks and Hispanics with a history of incarceration would be at even greater risk of food insecurity.

Finally, we explored factors that might mediate the association between history of incarceration and food insecurity. Several studies demonstrate that poverty is a predictor of food insecurity. According to the U.S. Department of Agriculture, about 29% of households under 185% of the federal poverty level were found to be food insecure in 2020, compared to only about 5% of households with incomes at 185% of the federal poverty level and above (Coleman-Jensen et al., 2021). Likewise, studies by Goldberg & Mawn (2015) and Morrissey et al. (2016) found that as poverty level increases, the likelihood of experiencing food insecurity also increases. Employment status, intricately linked to income, has also been associated with food insecurity. Coleman-Jensen (2011) found that households with heads who were unemployed, working part-time, or held multiple jobs, had greater odds of being food insecure than households with heads working one full-time job. Likewise, a multinational study conducted by Reeves et al. (2021) found a high predicted probability for moderate to severe food insecurity among the unemployed. Similarly, Huang et al. (2015) found that those who were unemployed had 55% increased odds of being food insecure. Given these known linkages between poverty, unemployment, and food insecurity, we tested the assumption that income level and employment status would mediate the association between history of incarceration and food insecurity. To address our research questions, we used cross-sectional data from the Health and Retirement Study, a population-based survey of community-dwelling U.S. older adults aged >50.

## Methods

### Study Design

This is a cross-sectional analysis pooling de-identified, publicly available survey data from the 2012 and 2014 waves of the Health and Retirement Study (HRS), an ongoing national longitudinal study of adults 51 and older (Heeringa & Connor, 1995). The HRS uses a four-stage probability sampling design to recruit adults aged 51 and older into the longitudinal study, oversampling non-Hispanic Blacks, Hispanics, and Florida residents, details of which are described elsewhere (Heeringa & Connor, 1995). Spouses of selected individuals are enrolled as well, regardless of age. During each wave of the HRS, surveyors collect data on a variety of demographic, lifestyle, health, biological, psychosocial, and physical measures. This current study was exempt from Michigan State University IRB review (STUDY00006614).

### Assessment of Incarceration History

History of incarceration was assessed via self-report in the 2012 and 2014 waves of the HRS. The item measuring history of incarceration was included in the psychosocial leave behind questionnaire, which is administered to a random 50% of all HRS respondents. Fifty percent of HRS respondents received the psychosocial leave behind questionnaire in 2012 and the other 50% of respondents received the questionnaire in 2014. Participants were asked if they had previously been inmates in a jail or prison (yes/no).

### Covariates

We adjusted for the following variables in our analyses: marital status (married, partnered, separated or divorced, widowed, never married), race/ethnicity (non-Hispanic White, non-Hispanic Black, non-Hispanic Other, Hispanic), gender (male/female), age (continuous variable), income (continuous variable), labor force status (working full-time, working part-time, unemployed, partly retired, retired, disabled, not in labor force), education (less than high school, GED, high school graduate, some college, college and above), and mother’s years of education (continuous variable). All categorical variables were dummy coded.

### Assessment of Food Insecurity

During the 2012 and 2014 waves of the HRS, one member of each household was asked the following question on behalf of the entire household: *“In the last two years, did you ever not have enough money to buy the food you need (yes/no)?”* Households were considered food insecure if the household respondent responded “yes” to this question.

### Sample Size

A total of 14,460 individuals completed the 2012 and 2014 HRS psychosocial leave behind questionnaires. We excluded 415 individuals who were 50 years of age and younger, another 104 individuals who did not respond to the questions about food insecurity and history of incarceration, and an additional 1,239 individuals missing data on any of the covariates. The final sample included 12,702 participants ages 51 and older.

### Analysis

We used multiple logistic regression to evaluate the association between history of incarceration and household food insecurity, adjusting for age, race/ethnicity, gender, education, marital status, employment status, and mother’s years of education. Associations were assessed using odds ratios (ORs) and 95% confidence intervals (CIs).

To evaluate differences in the association between incarceration and food insecurity based on race/ethnicity, we performed stratified analyses by racial/ethnic group. We calculated ORs and 95% CIs, adjusting for age, education, gender, marital status, and mother’s years of education. To test whether income level and/or employment status mediated the association between history of incarceration, we evaluated the association between history of incarceration and income using linear regression and between history of incarceration and employment using chi-squared analysis. We added each variable separately to our main logistical regression model evaluating the association between history of incarceration and food insecurity, adjusting for our standard covariates. Variables were considered potential mediators if they significantly reduced the association between history of incarceration and food insecurity in our model. All quantitative analyses were conducted using IBM SPSS Statistics (IBM, n.d.).

## Results

Our final sample included 12,702 respondents. The mean age of the sample was 68.4 (SD 10.3) and 59% of respondents were female (Table 1). Of the total sample, 71.1% of respondents were non-Hispanic White, 15.2% were non-Hispanic Black, 10.9% were Hispanic and 2.8% were of other racial/ethnic backgrounds. Sixty percent of respondents were married, and 85.8% had at least a high school education or equivalent. Fifty-seven percent of respondents were retired, and twenty-three percent were working full-time. Of the total sample, 12.8% reported food insecurity.

**Table 1.** Characteristics of Study Population (n = 12,702)

| Variable | Overall | History of Incarceration |  | p-value |
| --- | --- | --- | --- | --- |
|  |  | Yes | No |  |
| **% Food Insecure** | 12.8 | 27.6 | 11.7 | <.001 |
| **Gender** |  |  |  | <.001 |
| % Female | 59.4 | 21.5 | 62.2 |  |
| % Male | 40.6 | 78.5 | 37.8 |  |
| **Race/Ethnicity** |  |  |  | <.001 |
| % non-Hispanic White | 71.1 | 53.8 | 72.4 |  |
| % non-Hispanic Black | 15.2 | 28.6 | 14.2 |  |
| % non-Hispanic Other | 2.8 | 3.8 | 2.8 |  |
| % Hispanic | 10.9 | 13.9 | 10.7 |  |
| **Marital Status** |  |  |  | <.001 |
| % Married | 60.2 | 51.2 | 60.8 |  |
| % Partnered | 4.6 | 13.0 | 3.9 |  |
| % Separated or Divorced | 13.1 | 21.4 | 12.5 |  |
| % Widowed | 17.8 | 6.9 | 18.6 |  |
| % Never Married | 4.4 | 7.5 | 4.1 |  |
| **Education** |  |  |  | <.001 |
| % Less Than High School | 14.2 | 21.6 | 13.6 |  |
| % GED | 4.8 | 11.3 | 4.4 |  |
| % High School Graduate | 29.5 | 23.4 | 29.9 |  |
| % Some College | 26.1 | 29.2 | 25.9 |  |
| % College and Above | 25.4 | 14.4 | 26.2 |  |
| **Employment Status** |  |  |  | <.001 |
| % Works Full-Time | 22.9 | 26.6 | 22.6 |  |
| % Works Part-Time | 4.9 | 5.2 | 4.9 |  |
| % Unemployed | 2.3 | 5.5 | 2.1 |  |
| % Partially Retired | 8.4 | 6.8 | 8.5 |  |
| % Retired | 56.7 | 50.9 | 57.1 |  |
| % Disabled | 1.5 | 3.1 | 1.4 |  |
| % Not in Labor Force | 3.3 | 1.9 | 3.4 |  |
| **Mean Mother's Years Education (SD)** | 10.0 (3.7) | 9.8 (3.9) | 10.0 (3.7) | .088 |
| **Mean Age, Years (SD)** | 68.4 (10.3) | 63.4 (8.7) | 68.8 (10.3) | <.001 |
| **Mean Annual Income, U.S. Dollars (SD)** | 16,206.50 (41,632.80) | 13,693.29 (29,513.04) | 16,393.35 (42,390.70) | .064 |

When entered into the logistic regression model simultaneously, the following covariates were related to *increased* risk of food insecurity: female gender (OR=1.25; 95% CI: 1.10-1.42); non-Hispanic Black race/ethnicity (compared to non-Hispanic Whites; (OR=2.56; 95% CI: 2.23-2.94)); Hispanic ethnicity (OR=1.94; 95% CI: 1.62-2.34); other racial/ethnic group status (OR=1.74; 95% CI: 1.28-2.36); being partnered (compared to being married (OR=1.97; 95% CI: 1.56-2.48)); being separated or divorced (OR=2.56; 95% CI: 2.21-2.97); being widowed (OR=1.97; 95% CI: 1.65- 2.34); never being married (OR=2.38; 95% CI: 1.90-2.99); having left high school (compared to those with at least a college degree; (OR=2.98; 95% CI: 2.41-3.67)); having a GED (OR=2.53; 95% CI: 1.94-3.28); being a high school graduate (OR=2.01; 95% CI: 1.68-2.41); and having some college (OR=1.56; 95% CI: 1.25-1.81). The following covariates were associated with *decreased* risk of food insecurity: older age (OR=0.94; 95% CI: 0.94-0.95) and mother’s years of education (OR= 0.97; 95% CI: 0.95-0.99).

Seven percent (n=879) of respondents reported a history of incarceration. In a logistic regression model including all the covariates, we found that history of incarceration was associated with increased odds of food insecurity. After adjusting for education, gender, race/ethnicity, marital status, mother’s educational years, and age, those with a history of incarceration showed 83% increased odds (OR 1.83; 95% CI 1.52-2.21) of being food insecure compared to those without a history of incarceration (Figure 1).

**Figure 1:**
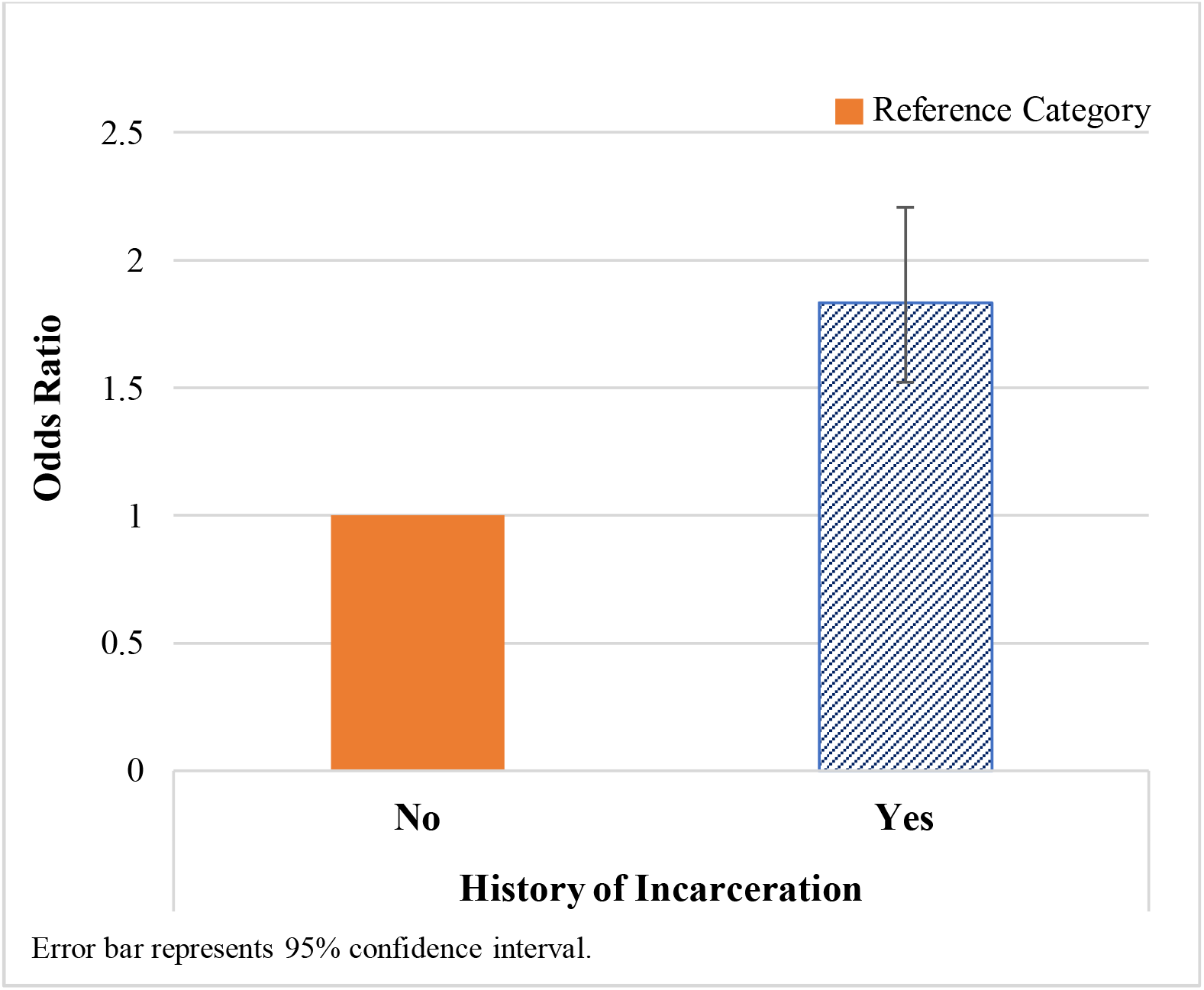
Odds of Reporting Food Insecurity

Separate logistic regression models were constructed to assess food insecurity among each racial/ethnic group, comparing those with a history of incarceration to those without a history of incarceration. All models were adjusted for education, gender, marital status, mother’s educational years, and age. Compared to non-Hispanic Whites without a history of incarceration, non-Hispanic Whites with a history of incarceration had 127% increased odds (OR 2.27; 95% CI 1.74-2.97) of food insecurity (Figure 2). Compared to non-Hispanic Blacks without a history of incarceration, non-Hispanic Blacks with a history of incarceration had 78% increased odds (OR 1.78; 95% CI 1.29-2.46) of food insecurity. There was no significant association between history of incarceration and food insecurity among Hispanics (OR 1.11; 95% CI 0.69-1.77) or non-Hispanics of other backgrounds (OR 1.79; 95% CI 0.71-4.52).

**Figure 2:**
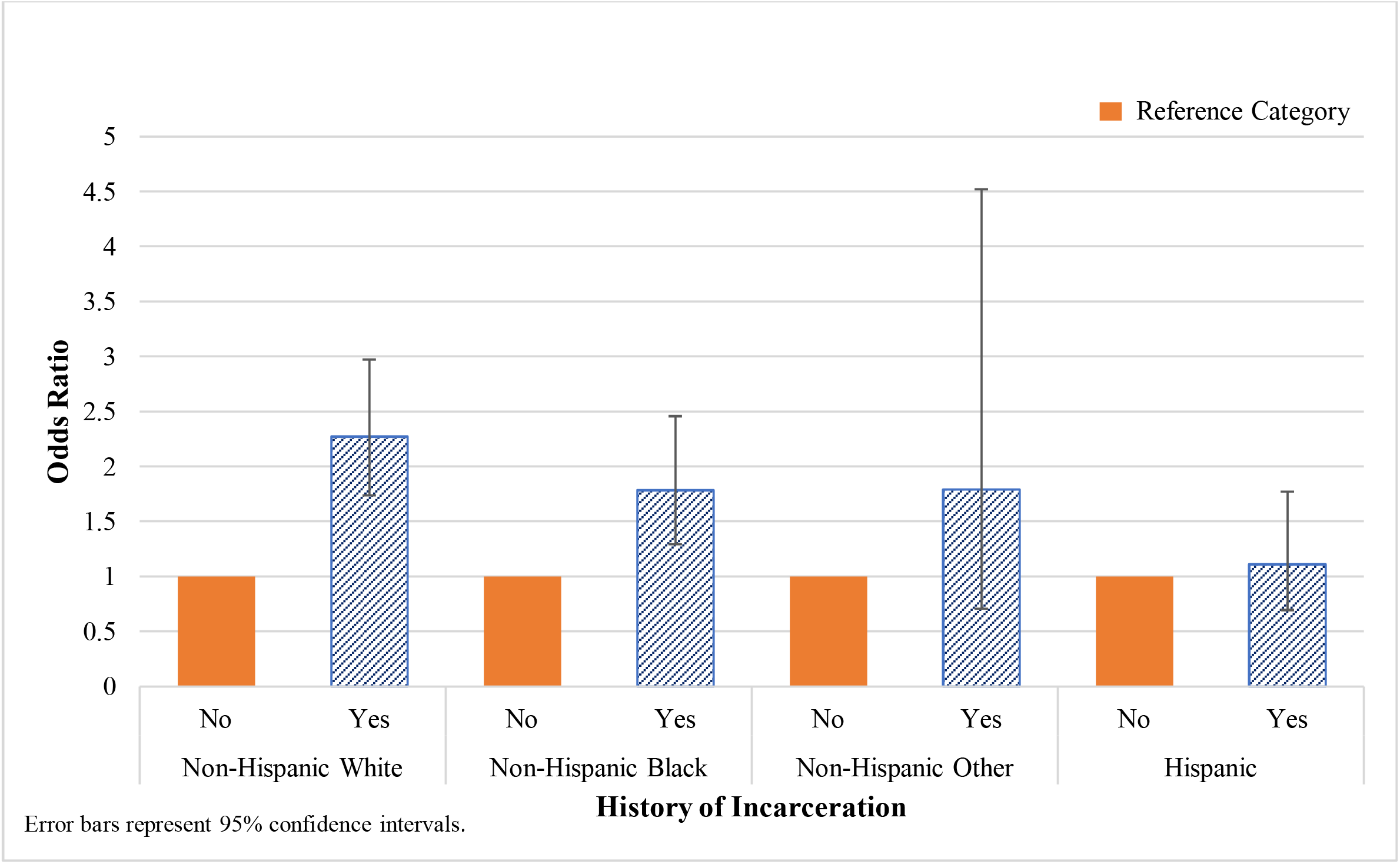
Odds of Reporting Food Insecurity by Race/Ethnicity and History of Incarceration

Finally, we were interested in whether respondent employment status and income mediated the association between history of incarceration and food insecurity. We observed that history of incarceration was associated with both income (β = -.038; p < .001) and employment status (□ ^2^ (6, N = 12702) = 77.16, p <.001). However, the association between history of incarceration and food insecurity was not reduced when adding income or employment status to our logistic regression model.

## Discussion

This study examined the association between history of incarceration and household food insecurity among U.S. community-dwelling older adults. We found that older adults with a history of incarceration had 83% increased odds of household food insecurity compared to those without a history of incarceration. This was true even after adjusting for potential confounding factors. The findings from this study are consistent with findings from studies of households with young children (Cox & Wallace, 2016), recently incarcerated new parents (Testa & Fahmy, 2021), and young adults (Testa & Jackson, 2019).

We also evaluated whether race/ethnicity moderated the association between history of incarceration and food insecurity. We found that there was an association between history of incarceration and food insecurity among non-Hispanic Whites and non-Hispanic Blacks, but not among Hispanics or those of other racial backgrounds. One possible explanation is that Hispanics and non-Hispanics of other racial groups tend to have larger household sizes than non-Hispanic Whites and non-Hispanic Blacks (U.S. Census Bureau, 2016). More importantly, Hispanics and non-Hispanics of other racial groups tend to have more people ages 18+ within the household (U.S. Census Bureau, 2016). Having more adults in the household may increase household economic resources in ways that decrease risk of food insecurity among Hispanics and other non-Hispanic groups.

It is also possible that racial/ethnic differences in food sources account for differences in food insecurity. Hispanic families are more likely than non-Hispanics to utilize WIC program services (Quandt et al., 2004; Sharkey et al., 2013). Non-Hispanic Blacks (Zekeri, 2007) and Hispanics (Kaufman & Karpati, 2007) report stigma as a factor when considering enrollment in food stamps programs. However, this does not seem to be a true barrier to enrollment for Hispanics (Kaufman & Karpati, 2007). Hispanics also rely heavily on social networks to cope with food insecurity, such as grandmothers and relationships with local bodega owners (Kaufman & Karpati, 2007). Given the limitations of the HRS data, we were unable to test these hypotheses. Future studies should consider how enrollment in supplemental food programs and social networks might mediate the association between incarceration history and food insecurity.

Additionally, differences in the amount of time spent incarcerated may explain racial/ethnic differences in the association between food insecurity and incarceration. In a previous study, Jordan & Freiburger (2015) found that Hispanics receive shorter sentences than non-Hispanic Whites and non-Hispanic Blacks. Additionally, Hispanics have been found to be released on parole more quickly than non-Hispanic Whites and non-Hispanic Blacks (Huebner & Bynum, 2008). To the authors’ knowledge, no studies have been conducted to identify the association between length of incarceration and food insecurity. Shorter length of incarceration may increase prospects for employment in ways that reduce food insecurity. Due to lack of efficient data on length of incarceration available in the HRS, we were unable to evaluate this association in the current study. Future studies should focus on identifying this association between length of incarceration and food insecurity.

Based on previous studies identifying an association between income and employment and food insecurity, we also evaluated whether income and employment mediated the relationship between history of incarceration and food insecurity. Although both income and employment status were associated with incarceration, neither of them reduced the association between history of incarceration and food insecurity. Future studies should explore other factors that might mediate the association between prior incarceration and food insecurity.

This study has several strengths. First, it utilized a large population-based sample of community-dwelling older adults with an oversampling of non-Hispanic Blacks and Hispanics. This is important, as previous studies have demonstrated that non-Hispanic Blacks and Hispanics experience high rates of incarceration (Carson, 2020). However, many studies regarding food insecurity among older adults tend to include fewer non-White participants (Afulani et al., 2015) or do not delineate race/ethnicity at all (Grammatikopoulou et al., 2019). Additionally, the HRS includes comprehensive demographic and economic data, which allowed us to explore these factors as other possible determinants of food insecurity.

This study is not without limitations. Due to the cross-sectional nature of this study, causality cannot be inferred. Additionally, we did not have data on length of incarceration, which may moderate the association between incarceration and food insecurity or explain observed racial ethnic/differences. Also, due to the secondary nature of this study, variables were limited to what was available from the primary data. Despite these limitations, to the authors’ knowledge, this is the first study that describes the association between food insecurity and history of incarceration using a population-based sample of older adults.

Our work has important implications for health policy and health promotion activities. There are numerous food assistance programs available to community-dwelling older adults, including The Supplemental Nutrition Assistance Program (SNAP; Food and Nutrition Service, n.d.a), Meals on Wheels (Meals on Wheels America, n.d.), The Commodity Supplemental Food Program (Food and Nutrition Service, n.d.b), The Senior Farmers’ Market Nutrition Program (Food and Nutrition Service, n.d.c), and The Emergency Food Assistance Program (Food and Nutrition, n.d.d). In some states, however, some of these programs have eligibility restrictions that may exclude certain individuals (i.e., those with felony drug convictions) from participation (Wolkomir, 2018). Current and future food assistance social service programs and interventions should be developed and implemented to include older adults with a history of incarceration.

Steps should also be taken to ensure that formerly incarcerated older adults are being connected with these programs as well. Additionally, due to current policy restrictions, formerly incarcerated individuals face a plethora of collateral consequences that may also contribute to food insecurity, such as difficulty finding stable employment (Welsh, 2015) and stable housing (Geller & Curtis, 2011). System policies such as these should be re-examined and implemented such that this population is no longer being excluded. Future studies should be conducted to build upon the results of this study as well as to further contribute to the current literature that is sorely lacking in this subject area.

## Data Availability

All data are available online at: https://hrsdata.isr.umich.edu/data-products/2012-hrs-core, https://hrsdata.isr.umich.edu/data-products/2014-hrs-core, and https://hrsdata.isr.umich.edu/data-products/rand-hrs-longitudinal-file-2018

## Acknowledgments

This analysis uses data from the Health and Retirement Study (2012 HRS Core, 2014 HRS Core, and RAND HRS Longitudinal File 2018), sponsored by the National Institute on Aging (grant number NIA U01AG009740) and conducted by the University of Michigan.

